# Cerebellar Network Compensation in Parkinson’s Disease: Functional Connectivity Across Motor and Cognitive Circuits

**DOI:** 10.64898/2026.05.19.26352927

**Authors:** Chi-Ying R. Lin, Thamires N.C. Magalhães, Shayla S. Yonce, Ihika Rampalli, Rory Mahabir, Jessica A. Bernard, the Parkinson’s Progression Markers Initiative (PPMI)

## Abstract

**Introduction.:** The cerebellum is increasingly recognized as a key contributor to cognitive reserve and network adaptation in Parkinson’s disease (PD). However, how cerebello–cortical and cerebello–basal ganglia connectivity reorganizes across disease duration and cognitive status remains incompletely understood.

**Methods.:** Resting-state fMRI data from the Parkinson’s Progression Markers Initiative were analyzed in 172 individuals with PD. We investigated cerebello-basal ganglia and cerebello-cortical connectivity using ROI-to-ROI and seed-to-voxel pipelines respectively, providing novel insights into both subcortical and cortical effects. Effects of age, disease duration, cognitive status, motor symptom severity, and dopaminergic medication were assessed.

**Results.:** Across all participants, cerebellar lobule VI and vermis VI showed robust positive connectivity with the pallidum, along with high intra-cerebellar coupling. When controlling for dopaminergic medication, lobule V connectivity with the primary motor cortex was reduced. Age was associated with lower cerebello–basal ganglia connectivity widespread across nodes, evident across medication states. Disease duration showed region-specific effects: in cognitively normal PD, longer duration corresponded to stronger lobule V–temporal cortex connectivity and higher Crus I–precentral gyrus connectivity than PD with cognitive dysfunction. Motor symptom severity was not related to connectivity.

**Conclusions.:** Cerebellar connectivity patterns in PD are linked to disease duration and cognitive preservation. Enhanced cerebello–cortical coupling in cognitively normal PD may reflect compensatory network recruitment that diminishes with cognitive decline. These findings position cerebellar networks as candidate markers of compensatory capacity and disease trajectory in PD.

## Introduction

Parkinson’s disease (PD) is the second most common neurodegenerative disorder, affecting more than 10 million people worldwide.^1^ It is traditionally defined by its hallmark motor symptoms, including bradykinesia, rigidity, tremor, and postural instability, resulting from dopaminergic cell loss in the substantia nigra pars compacta. However, the disease extends far beyond motor dysfunction: cognitive impairment is a particularly distressing non-motor feature, with cumulative prevalence up to 50% of PD individuals eventually developing dementia within 10 years of the diagnosis and 80% after 20 year of diagnosis.^2^ Importantly, the trajectory from cognitively normal PD to mild cognitive impairment and dementia is highly variable,^3^ suggesting that compensatory mechanisms and brain reserve play a significant role in modulating the manifestations of non-motor features of PD.

Despite accounting for only close to 10% of intracranial volume, the cerebellum houses the majority of the brain’s neurons.^4–6^ Over the past several decades, the cerebellum been recognized as an important hub to maintain cognitive function, supporting executive function, language, and working memory, in addition to its classically recognized contributions to motor behavior.^7,8^ Additionally, the cerebellar regions that are most strongly implicated in cognition also maintain robust connections with motor and basal ganglia circuits, positioning the “cognitive cerebellum” as an interface between motor and higher-order networks in PD.^9^ Prior studies exhibited significant cerebellar network alterations in Parkinson’s disease and identifies their co-occurrence with cerebellar atrophy.^9–13^ Additionally, non-linear progression of cerebellar microstructural changes in PD was identified, reflecting adaptive reorganization concurrent with ongoing striatal dopaminergic decline.^14^ Importantly, mirroring observations in Alzheimer’s disease, the cerebellum is largely preserved from direct early PD pathology, retaining its structural integrity and showing enhanced or reorganized functional connectivity.^15,16^ Such preservation implies that the cerebellum may provide a compensatory substrate that supports *network-level adaptation* across the trajectory of PD.^9,17–19^ These findings overall support the hypothesis that the cerebellum can contribute to neural scaffolding and reserve in aging and neurodegenerative diseases.^7,20–24^ Despite this, the role of cerebellar network changes in PD disease trajectory remains underexplored.

In the present study, we investigate whether cerebellar functional connectivity provides compensatory support for cognition and motor-related networks across the spectrum of PD. Using resting-state fMRI from the Michael J. Fox Foundation Parkinson’s Progression Markers Initiative,^16^ we examine the functional connectivity between cerebellar lobules and both cerebral cortical and subcortical regions. In particular, we focused on subcortical connections between the cerebellum and basal ganglia, given the role of these circuits in PD pathophysiology. Additionally, prior work in aging has demonstrated functionally segregated topographic motor and cognitive cerebellar-basal ganglia circuits, which are relevant for behavior and impacted in advanced age, likely reflecting age-related dopamine decline.^25^ These same circuits are modulated by dopaminergic manipulation in both healthy adults and individuals with PD, underscoring their clinical relevance across disease progression.^10,26^ We hypothesized that cerebellar functional connectivity with cortical and basal ganglia networks is altered in PD, reflecting compensatory responses to dopaminergic decline acting on age-related circuit vulnerability.

## Methods

Data used in the preparation of this article was obtained on January 2, 2025 from the Parkinson’s Progression Markers Initiative (PPMI) database (www.ppmi-info.org/access-data-specimens/download-data), RRID:SCR_006431.^27^ For up-to-date information on the study, visit www.ppmi-info.org. This analysis used data openly available from PPMI. Demographic data was selected from the “Participant Status, Demographics, Socio-Economic, and PD Diagnosis History” files. Entries were selected by filtering for patients designated as controls and having PD per the PD Diagnosis History file. For each patient, baseline data was selected. Only participants with MRI and fMRI were included in the study. Cerebellar coverage was evaluated by two trained lab members (TM and MM) and independently verified by the senior author (JAB). Participants were excluded if cerebellar imaging did not extend below Crus I. A total of 172 PD participants were then included. Demographic and clinical covariate data included age, sex, education (years), disease duration (years), Montreal Cognitive Assessment (MoCA) score, International Parkinson’s and Movement Disorders Society Unified Parkinson’s Disease Rating Scale (MDS-UPDRS) part III score, and PD medication history. The dopaminergic medication was stratified into three categories for statistical investigation: i) carbidopa/levodopa (regular or extended release), ii) non-carbidopa/levodopa dopaminergic medication (amantadine, selegiline, rasagiline, ropinirole, pramipexole), and iii) drug naïve. With a total score of 132, the MDS-UPDRS was stratified into mild (32 ≤ below), moderate (33-58), and severe (≥ 59) to represent different level of motor impairment severity. Furthermore, from the cognitive status point of view, MoCA > 26 was categorized as PD with normal cognitive function, and MoCA < 26 is PD with cognitive dysfunction.^28^

### Neuroimaging Data Processing and Analysis

All data processing followed procedures used in our recent work.^29^ For the sake of clarity, consistency, and replication, we have restated those here using boilerplate text with additional details about choices made that were specific to this data set. All data were pre-processed using the CONN toolbox (22.v2407).^30^ Functional and anatomical data were preprocessed using a flexible preprocessing pipeline including realignment, outlier detection, direct segmentation and MNI-space normalization, and smoothing.^31^ Functional data were realigned using the SPM realign & unwarp procedure,^32^ where all scans were co-registered to a reference image (first scan of the first session) using a least squares approach and a 6 parameter (rigid body) transformation,^33^ and resampled using b-spline interpolation. However, we did not incorporate slice timing correction given the nature of the data analyzed here. Because this data set includes scans collected across multiple sites, the inclusion of re-slicing during realignment could result in inconsistencies and errors in processing, and this allows for more reliable motion parameter estimation. This also avoids challenges and inaccuracies that would result from incorrect slice-order assumptions. Potential outlier scans were identified using ART as acquisitions with subject motion above 0.9 mm or global BOLD signal changes above 5 standard deviations,^30,31,34^ which is the equivalent of the “intermediate” settings as defined by CONN. A reference BOLD image was computed for each subject by averaging all scans excluding outliers. Functional and anatomical data were normalized into standard MNI space, segmented into grey matter, white matter, and CSF tissue classes, and resampled to 2 mm isotropic voxels following a direct normalization procedure using SPM unified segmentation and normalization algorithm with the default tissue probability map template.^31,35–37^ Last, functional data were smoothed using spatial convolution with a Gaussian kernel of 8 mm full width half maximum (FWHM).

Functional data were denoised using a standard denoising pipeline,^31^ including the regression of potential confounding effects characterized by white matter timeseries (5 CompCor noise components), CSF timeseries (5 CompCor noise components), motion parameters and their first order derivatives (12 factors),^38^ outlier scans (12 factors),^34^ and linear trends (2 factors) within each functional run, followed by bandpass frequency filtering of the BOLD timeseries between 0.008 Hz and 0.09 Hz.^39^ This was applied to the resting state data in order to focus on slow-frequency fluctuations while minimizing the influence of physiological, head-motion and other noise sources.^39^ Potential confounding effects to the estimated BOLD signal were determined and removed separately for each voxel and for each subject and functional run/session, using Ordinary Least Squares (OLS) regression to project each BOLD signal timeseries to the sub-space orthogonal to all potential confounding effects using an anatomical component-based noise correction procedure (aCompCor).^40^

All statistical analyses of resting-state functional connectivity were also completed in CONN. Data was analyzed using a targeted ROI-to-ROI approach in order to focus on the cerebello-basal ganglia circuit. This was then followed by exploratory whole brain analyses of our cerebellar regions of interest to better understand differences in cerebellar-whole brain connectivity associated with cognitive decline in PD. For ROI-to-ROI analyses, cerebellar regions included bilateral hemispheric lobules I–IV, V, VI, Crus I, Crus II, and corresponding vermal regions (vermis VI, vermis Crus I, and vermis Crus II). All regions were as defined by the SUIT atlas. Subcortical ROIs included bilateral caudate, putamen, pallidum, and accumbens using the atlas-based ROIs available in CONN.^41,42^ We first established patterns of cerebello-basal ganglia connectivity in all PD patients, and further investigated impacts of age, disease duration, and motor symptom severity as indexed by the MDS-UPDRS. Connectivity results were considered significant at a connection-level threshold of *p < 0.05* and a cluster-level threshold of *pFDR < 0.05* (MVPA omnibus test). For seed-to-voxel analyses, we focused on Lobule V and Crus I in the right hemisphere to reduce multiple comparison burden and to serve as a targeted initial examination of cerebellar connectivity patterns as we explored differences in those with and without cognitive decline. Prior work looking at both the left and right cerebellar hemispheres has demonstrated comparable patterns of connectivity in these seeds;^43–45^ as such, we chose to focus on the right hemisphere. All whole-brain analyses were evaluated using threshold-free cluster enhancement (TFCE),^46^ a robust statistical approach implemented using 1,000 permutations. Like in the ROI-ROI analyses, we also investigated effects of disease duration and motor symptom severity. These seed-based analyses were implemented as supplementary validation steps to confirm the expected network topology and connectivity patterns.

## Results

The 172 participants with PD had an average age of 65.0 ± 7.8 years, including 60 women and 112 men. They had 16.6 ± 3.7 years of education, a MoCA score of 26.9 ± 2.4, and a MDS-UPDRS Part III score of 22.1 ± 10.3. Disease duration was 49.3 ± 37.6 months. Based on MoCA scores, 105 participants were cognitively normal (MoCA ≥ 26), whereas 67 showed cognitive dysfunction (MoCA < 26; 58 with MoCA 23-26, and 9 with MoCA ≤ 22). With respect to medication status, 68 participants were treated with carbidopa/levodopa, 64 received non-carbidopa/levodopa dopaminergic medications, and 40 were medication-naïve. Disease severity, assessed using the Hoehn and Yahr (H&Y) scale, was distributed as follows: 62 participants were in stage 1, 58 in stage 2, and 52 in stage 3.

### Foundational cerebellar network alterations in PD

We first asked whether foundational cerebellar network alterations could be detected in PD regardless of cognitive function and while controlling for dopaminergic medication (**Figure 1**). Using our ROI-ROI approach to investigate cerebellar-basal ganglia connectivity, we found patterns of both positive and negative connectivity (anti-correlations) between cerebellar and basal ganglia regions. The complete results are detailed in **Table 1**. Of note, across all PD participants, robust positive connectivity was identified between the globus pallidus and both right cerebellar lobule VI (lobule VI – pallidum) and vermis VI (vermis VI – pallidum) (**Figure 1A**). Lobules I-IV, V, and VI also showed positive associations with the putamen, reflecting the established motor (i.e., lobule I-V) and cognitive (i.e., lobule V and VI) dissociation within the cerebello-striatal circuit, as well as robust intra-cerebellar connections (**Table 1**, **Figure 1A**). These results suggest that cerebellar circuits may be upregulated to compensate for basal ganglia dysfunction in PD. When investigating whole brain connectivity, Lobule V demonstrated the expected canonical pattern of positive correlations with motor cortical regions and anti-correlations with frontal and association areas, supporting the validity of the connectivity pipeline (**Table 2**, **Figure 1B**).^47–49^ A comparable connectivity pattern was observed for Crus I, reflecting its well-established canonical role within the frontal association network. However, when controlling for dopaminergic medication, Lobule V connectivity with primary motor cortex (M1) was no longer present across all PD participants (**Table 2**, **Figure 1C**), and there were no group differences identified when comparing cognitively normal PD participants to those with cognitive dysfunction. This suggests that medication may modulate cerebello-cerebral motor coupling, regardless of cognitive capacity. There were no subgroup differences observed for Crus I, and medication effects were less pronounced relative to Lobule V. The overall connectivity pattern of canonical Crus I network remained intact, though less extensive.

**Figure 1.**
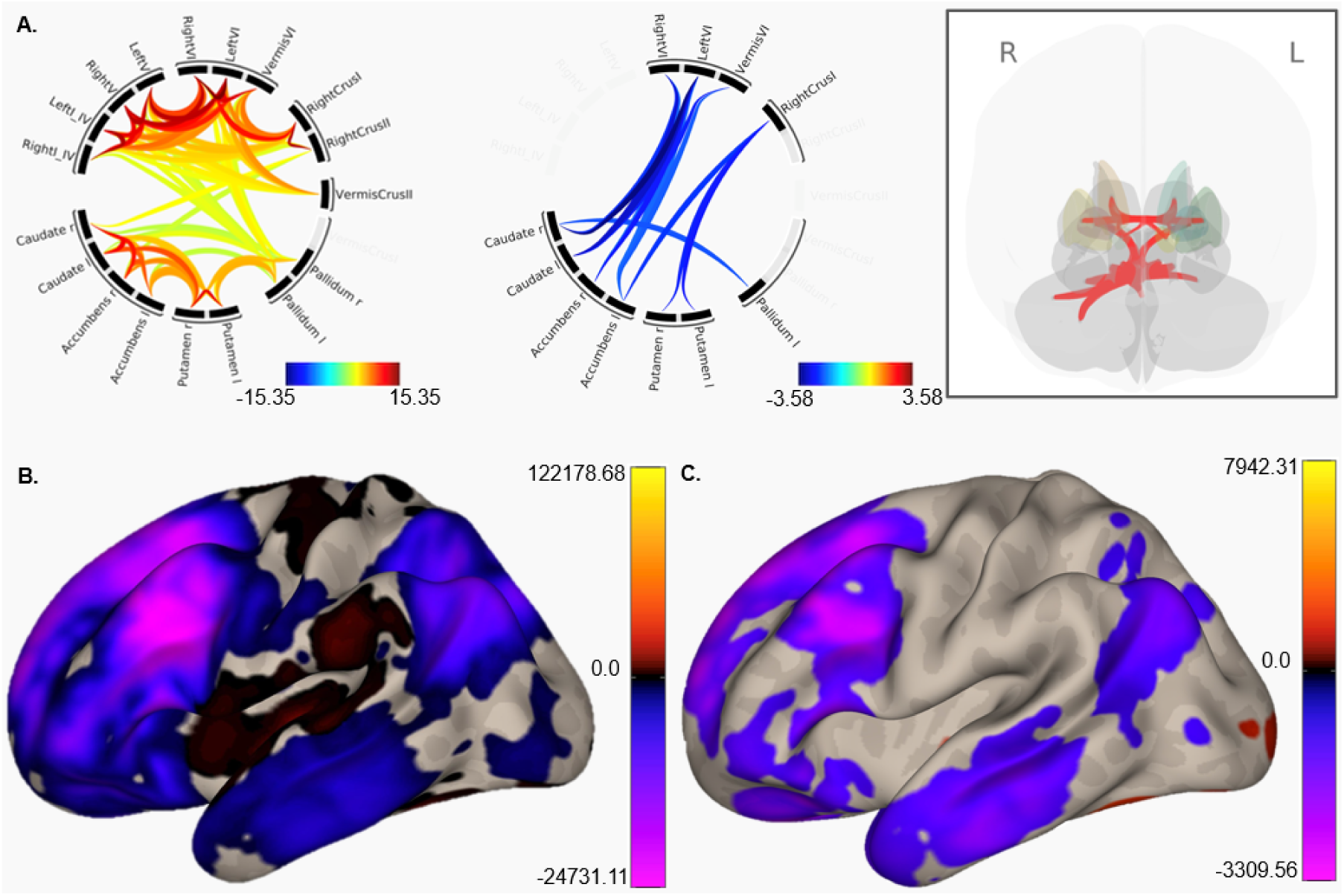
Foundational cerebellar network alterations in Parkinson’s disease. A. Connectome rings showing the cerebello-basal ganglia circuits in all PD patients, while controlling for dopaminergic medication. The hot colors (left) represent positive correlations between ROIs while the cool colors (right) show negative correlations. The inset (gray box) includes a 3D visualization of the positive associations between ROIs within and between the cerebellum and basal ganglia. Color bars indicate the strength of the relationships between ROIs. R: right; L: left. B. Connectivity between cerebellar Lobule V and the cortex in all PD patients demonstrating positive associations with motor and premotor cortical regions, and anti-correlations with frontal, temporal, and parietal cortices, consistent with past findings in healthy young and older adults. C. When controlling for dopaminergic medication, the pattern of Lobule V-cortical connectivity is lessened, and the associations with motor cortical regions are no longer significant. Chord diagrams depict the magnitude and direction of age-related associations. All results are thresholded at *p < 0.05* and cluster-level *pFDR < 0.05* (MVPA omnibus test). Color bars for B and C represent the significant connectivity patterns, as indexed by TFCE values.

**Table 1.** Region of interest (ROI) to ROI analyses for the cerebello-basal ganglia functional network across the full Parkinson’s disease cohort.

|  | Region 1 | Region 2 | Statistic | p-unc | p-FDR |
| --- | --- | --- | --- | --- | --- |
| Positive | Right VI | Left VI | T(170) = 15.35 | < 0.0001 | < 0.0001 |
|  | Vermis VI | Right VI | T(170) = 12.52 | < 0.0001 | < 0.0001 |
|  | Vermis VI | Left VI | T(170) = 11.99 | < 0.0001 | < 0.0001 |
|  | Right I to IV | Left I to IV | T(170) = 13.77 | < 0.0001 | < 0.0001 |
|  | Right V | Left V | T(170) = 13.61 | < 0.0001 | < 0.0001 |
|  | Left V | Left I to IV | T(170) = 12.14 | < 0.0001 | < 0.0001 |
|  | Right V | Right I to IV | T(170) = 12.05 | < 0.0001 | < 0.0001 |
|  | Right V | Left I to IV | T(170) = 10.70 | < 0.0001 | < 0.0001 |
|  | Left V | Right I to IV | T(170) = 10.37 | < 0.0001 | < 0.0001 |
|  | Right Crus I | Left Crus I | T(170) = 14.73 | < 0.0001 | < 0.0001 |
|  | Left Crus II | Left Crus I | T(170) = 14.31 | < 0.0001 | < 0.0001 |
|  | Right Crus II | Right Crus I | T(170) = 13.84 | < 0.0001 | < 0.0001 |
|  | Left Crus II | Right Crus II | T(170) = 12.67 | < 0.0001 | < 0.0001 |
|  | Left Crus II | Right Crus I | T(170) = 9.70 | < 0.0001 | < 0.0001 |
|  | Right Crus II | Left Crus I | T(170) = 9.56 | < 0.0001 | < 0.0001 |
|  | Right putamen | Left putamen | T(170) = 12.08 | < 0.0001 | < 0.0001 |
|  | Right caudate | Left caudate | T(170) = 11.79 | < 0.0001 | < 0.0001 |
|  | Right caudate | Right accumbens | T(170) = 10.75 | < 0.0001 | < 0.0001 |
|  | Left caudate | Right putamen | T(170) = 9.06 | < 0.0001 | < 0.0001 |
|  | Right accumbens | Left accumbens | T(170) = 8.99 | < 0.0001 | < 0.0001 |
|  | Left caudate | Left accumbens | T(170) = 8.66 | < 0.0001 | < 0.0001 |
|  | Left caudate | Right accumbens | T(170) = 7.93 | < 0.0001 | < 0.0001 |
|  | Right accumbens | Right putamen | T(170) = 7.31 | < 0.0001 | < 0.0001 |
|  | Left accumbens | Right putamen | T(170) = 6.61 | < 0.0001 | < 0.0001 |
|  | Left accumbens | Left putamen | T(170) = 6.12 | < 0.0001 | < 0.0001 |
|  | Right caudate | Left accumbens | T(170) = 6.20 | < 0.0001 | < 0.0001 |
|  | Left caudate | Left putamen | T(170) = 6.05 | < 0.0001 | < 0.0001 |
|  | Right caudate | Right putamen | T(170) = 5.51 | < 0.0001 | < 0.0001 |
|  | Right caudate | Left putamen | T(170) = 5.45 | < 0.0001 | < 0.0001 |
|  | Right accumbens | Left putamen | T(170) = 5.04 | < 0.0001 | < 0.0001 |
|  | Left VI | Left V | T(170) = 13.44 | < 0.0001 | < 0.0001 |
|  | Right VI | Right V | T(170) = 13.24 | < 0.0001 | < 0.0001 |
|  | Left VI | Right V | T(170) = 11.23 | < 0.0001 | < 0.0001 |
|  | Vermis VI | Right V | T(170) = 10.63 | < 0.0001 | < 0.0001 |
|  | Right VI | Left V | T(170) = 9.49 | < 0.0001 | < 0.0001 |
|  | Vermis VI | Left V | T(170) = 8.66 | < 0.0001 | < 0.0001 |
|  | Right VI | Right I to IV | T(170) = 8.29 | < 0.0001 | < 0.0001 |
|  | Left VI | Left I to IV | T(170) = 7.64 | < 0.0001 | < 0.0001 |
| Positive | Vermis VI | Right I to IV | T(170) = 7.22 | < 0.0001 | < 0.0001 |
|  | Left VI | Right I to IV | T(170) = 7.00 | < 0.0001 | < 0.0001 |
|  | Right VI | Left I to IV | T(170) = 6.83 | < 0.0001 | < 0.0001 |
|  | Vermis VI | Left I to IV | T(170) = 6.17 | < 0.0001 | < 0.0001 |
|  | Left Crus I | Left VI | T(170) = 12.83 | < 0.0001 | < 0.0001 |
|  | Right Crus I | Right VI | T(170) = 10.98 | < 0.0001 | < 0.0001 |
|  | Left Crus I | Right VI | T(170) = 9.22 | < 0.0001 | < 0.0001 |
|  | Left Crus I | Vermis VI | T(170) = 8.19 | < 0.0001 | < 0.0001 |
|  | Right Crus I | Vermis VI | T(170) = 7.88 | < 0.0001 | < 0.0001 |
|  | Right Crus I | Left VI | T(170) = 7.47 | < 0.0001 | < 0.0001 |
|  | Left Crus II | Left VI | T(170) = 6.16 | < 0.0001 | < 0.0001 |
|  | Left Crus II | Vermis VI | T(170) = 5.88 | < 0.0001 | < 0.0001 |
|  | Right Crus II | Vermis VI | T(170) = 4.69 | < 0.0001 | < 0.0001 |
|  | Right Crus II | Right VI | T(170) = 4.61 | < 0.0001 | < 0.0001 |
|  | Left Crus II | Right VI | T(170) = 3.22 | 0.0015 | 0.0043 |
|  | Right Crus II | Left VI | T(170) = 2.06 | 0.0405 | 0.0675 |
|  | Right putamen | Left pallidum | T(170) = 6.04 | < 0.0001 | < 0.0001 |
|  | Left putamen | Left pallidum | T(170) = 6.10 | < 0.0001 | < 0.0001 |
|  | Right putamen | Right pallidum | T(170) = 5.87 | < 0.0001 | < 0.0001 |
|  | Left putamen | Right pallidum | T(170) = 4.06 | < 0.0001 | 0.0001 |
|  | Left caudate | Left pallidum | T(170) = 3.28 | 0.0006 | 0.0020 |
|  | Left caudate | Right pallidum | T(170) = 2.06 | 0.0413 | 0.0688 |
|  | Left accumbens | Left pallidum | T(170) = 1.99 | 0.0483 | 0.0882 |
|  | Vermis Crus II | Vermis VI | T(170) = 6.90 | < 0.0001 | < 0.0001 |
|  | Vermis Crus II | Left VI | T(170) = 6.07 | < 0.0001 | < 0.0001 |
|  | Vermis Crus II | Right VI | T(170) = 4.97 | < 0.0001 | < 0.0001 |
|  | Left pallidum | Left VI | T(170) = 4.15 | < 0.0001 | < 0.0001 |
|  | Right pallidum | Right VI | T(170) = 3.69 | 0.0001 | < 0.0001 |
|  | Right pallidum | Left VI | T(170) = 3.59 | 0.0004 | 0.0003 |
|  | Left pallidum | Right VI | T(170) = 3.13 | 0.0020 | 0.0015 |
|  | Right pallidum | Vermis VI | T(170) = 2.83 | 0.0053 | 0.0176 |
|  | Left pallidum | Vermis VI | T(170) = 2.47 | 0.0143 | 0.0280 |
|  | Left Crus I | Left V | T(170) = 6.57 | < 0.0001 | < 0.0001 |
|  | Left Crus I | Right V | T(170) = 6.27 | < 0.0001 | < 0.0001 |
|  | Right Crus I | Right V | T(170) = 5.75 | < 0.0001 | < 0.0001 |
|  | Right Crus I | Left V | T(170) = 5.66 | < 0.0001 | < 0.0001 |
|  | Right Crus I | Left I to IV | T(170) = 5.17 | < 0.0001 | < 0.0001 |
|  | Right Crus I | Right I to IV | T(170) = 4.96 | < 0.0001 | < 0.0001 |
|  | Left Crus I | Left I to IV | T(170) = 4.86 | < 0.0001 | < 0.0001 |
|  | Left Crus I | Right I to IV | T(170) = 4.57 | < 0.0001 | < 0.0001 |
|  | Left Crus II | Left V | T(170) = 3.48 | 0.0006 | 0.0021 |
|  | Right Crus II | Right I to IV | T(170) = 3.05 | 0.0026 | 0.0068 |
|  | Right Crus II | Right V | T(170) = 3.04 | 0.0027 | 0.0068 |
|  | Left Crus II | Left I to IV | T(170) = 3.00 | 0.0032 | 0.0077 |
|  | Left Crus II | Right V | T(170) = 2.96 | 0.0034 | 0.0077 |
|  | Right Crus II | Left I to IV | T(170) = 2.66 | 0.0085 | 0.0190 |
| **Positive** | Left Crus II | Right I to IV | T(170) = 2.50 | 0.0132 | 0.0265 |
|  | Right Crus II | Left V | T(170) = 2.36 | 0.0196 | 0.0356 |
|  | Left pallidum | Right pallidum | T(170) = 5.36 | < 0.0001 | < 0.0001 |
|  | Vermis Crus II | Left V | T(170) = 4.74 | 0.000002 | < 0.0001 |
|  | Vermis Crus II | Right V | T(170) = 4.32 | 0.000013 | 0.0001 |
|  | Vermis Crus II | Left I to IV | T(170) = 4.16 | 0.000025 | 0.0001 |
|  | Vermis Crus II | Right I to IV | T(170) = 3.52 | 0.0006 | 0.0014 |
|  | Left pallidum | Left I to IV | T(170) = 3.18 | 0.0017 | 0.0057 |
|  | Left pallidum | Right I to IV | T(170) = 2.98 | 0.0033 | 0.0083 |
|  | Left pallidum | Right V | T(170) = 2.86 | 0.0048 | 0.0107 |
|  | Right pallidum | Right I to IV | T(170) = 2.74 | 0.0068 | 0.0196 |
|  | Left pallidum | Left V | T(170) = 2.45 | 0.0154 | 0.0280 |
|  | Right pallidum | Right V | T(170) = 2.55 | 0.0116 | 0.0290 |
|  | Vermis Crus II | Left Crus I | T(170) = 4.23 | < 0.0001 | 0.0001 |
|  | Vermis Crus II | Left Crus II | T(170) = 2.19 | 0.0300 | 0.0599 |
|  | Vermis Crus II | Right Crus I | T(170) = 2.01 | 0.0457 | 0.0703 |
|  | Right caudate | Right Crus II | T(170) = 3.85 | 0.0002 | 0.0006 |
|  | Right accumbens | Right Crus II | T(170) = 2.41 | 0.0170 | 0.0483 |
|  | Left putamen | Left VI | T(170) = 2.03 | 0.0436 | 0.0968 |
| **Negative** | Left accumbens | Vermis Crus II | T(170) = -2.40 | 0.0174 | 0.0497 |
|  | Left caudate | Vermis Crus II | T(170) = -2.10 | 0.0369 | 0.0671 |
|  | Right caudate | Left pallidum | T(170) = -2.10 | 0.0373 | 0.0746 |
|  | Right caudate | Vermis Crus II | T(170) = -2.04 | 0.0433 | 0.0787 |
|  | Right putamen | Left Crus I | T(170) = -2.88 | 0.0045 | 0.1115 |
|  | Left accumbens | Left Crus I | T(170) = -2.80 | 0.0057 | 0.0191 |
|  | Left Accumbens | Right Crus I | T(170) = -2.32 | 0.0215 | 0.0538 |
|  | Right putamen | Right Crus I | T(170) = -2.45 | 0.0154 | 0.0343 |
|  | Left putamen | Right Crus I | T(170) = -2.60 | 0.0102 | 0.0256 |
|  | Right putamen | Left Crus II | T(170) = -2.02 | 0.0452 | 0.0903 |
|  | Right caudate | Left VI | T(170) = -3.58 | 0.0004 | 0.0013 |
|  | Left caudate | Left VI | T(170) = -3.04 | 0.0028 | 0.0079 |
|  | Left caudate | Right VI | T(170) = -3.00 | 0.0031 | 0.0079 |
|  | Left caudate | Vermis VI | T(170) = -2.47 | 0.0146 | 0.0291 |
|  | Right caudate | Right VI | T(170) = -2.35 | 0.0199 | 0.0441 |
|  | Left accumbens | Vermis VI | T(170) = -2.06 | 0.0413 | 0.0882 |
|  | Right accumbens | Left VI | T(170) = -2.57 | 0.0109 | 0.0363 |
|  | Left caudate | Left V | T(170) = -2.56 | 0.0114 | 0.0253 |
|  | Right caudate | Left V | T(170) = -2.51 | 0.0130 | 0.0326 |
|  | Right accumbens | Left V | T(170) = -2.35 | 0.0200 | 0.0492 |
|  | Right accumbens | Right V | T(170) = -2.22 | 0.0279 | 0.0620 |

Connectivity results were considered significant at a connection-level threshold of $p < 0.05$ and a cluster-level threshold of $pFDR < 0.05$ (MVPA omnibus test).

**Table 2.** Lobule V whole brain connectivity in Parkinson’s disease: unadjusted and medication-adjusted models.

| Condition | Regions | Cluster<br>(x,y,z) | Size | Peaks | TFCE | peak *p-FDR* |
| --- | --- | --- | --- | --- | --- | --- |
| **Without**<br>**Medication** | Right Crus I | +30 -42 -32 | 58239 | 60 | 903802.83 | < 0.0001 |
|  | Medial/superior frontal gyrus | +06 +40 +42 | 92635 | 281 | 24913.36 | < 0.0001 |
|  | Left precenral gyrus | -26 -22 +72 | 2316 | 15 | 3058.82 | < 0.0001 |
|  | Left inferior temporal gyrus | -50 +00 -48 | 20 | 2 | 826.39 | 0.0016 |
| **Controlling for**<br>**Medication** | Right VI | +28 -40 -32 | 33256 | 63 | 73179.78 | < 0.0001 |
|  | Medial superior frontal gyrus | -02 +34 +44 | 28716 | 164 | 3365.14 | < 0.0001 |
|  | Left lateral occipital cortex | -38 -76 +38 | 2591 | 19 | 1591.62 | < 0.0001 |
|  | Right lateral occipital cortex | +46 -70 +42 | 3055 | 11 | 1431.61 | 0.0001 |
|  | Precuneus/posterior cingulate gyrus | +06 -52 +38 | 1381 | 8 | 1263.80 | 0.0004 |
|  | Right anterior temporal pole | +34 +10 -40 | 3152 | 29 | 1259.00 | 0.0004 |
|  | Cingulate gyrus, anterior division | +04 +04 +30 | 38 | 3 | 864.54 | 0.0024 |
|  | Right postcentral gyrus | +52 -24 +58 | 47 | 4 | 861.70 | 0.0024 |

Connectivity results were considered significant at a connection-level threshold of $p < 0.05$ and a cluster-level threshold of $pFDR < 0.05$ (MVPA omnibus test).

### Effect of age

We further studied whether the effect of age impacts cerebellar functional connectivity in PD, controlling for the effect of dopaminergic medication. We found that in the full PD cohort, age was negatively associated with widespread functional connectivity between cerebellar lobules and all regions of basal ganglia (**Table 3**, **Figure 2**), consistent with patterns seen in healthy older adults as well.^25^ There are consistent age-related anti-correlations involving the pallidum, putamen, and caudate, as well as cerebellar lobules V, VI, and Crus I/II. However, there were no statistically significant group differences between cognitively normal PD and PD with cognitive dysfunction in the relationships between cerebello-basal ganglia connectivity and age.

**Figure 2.**
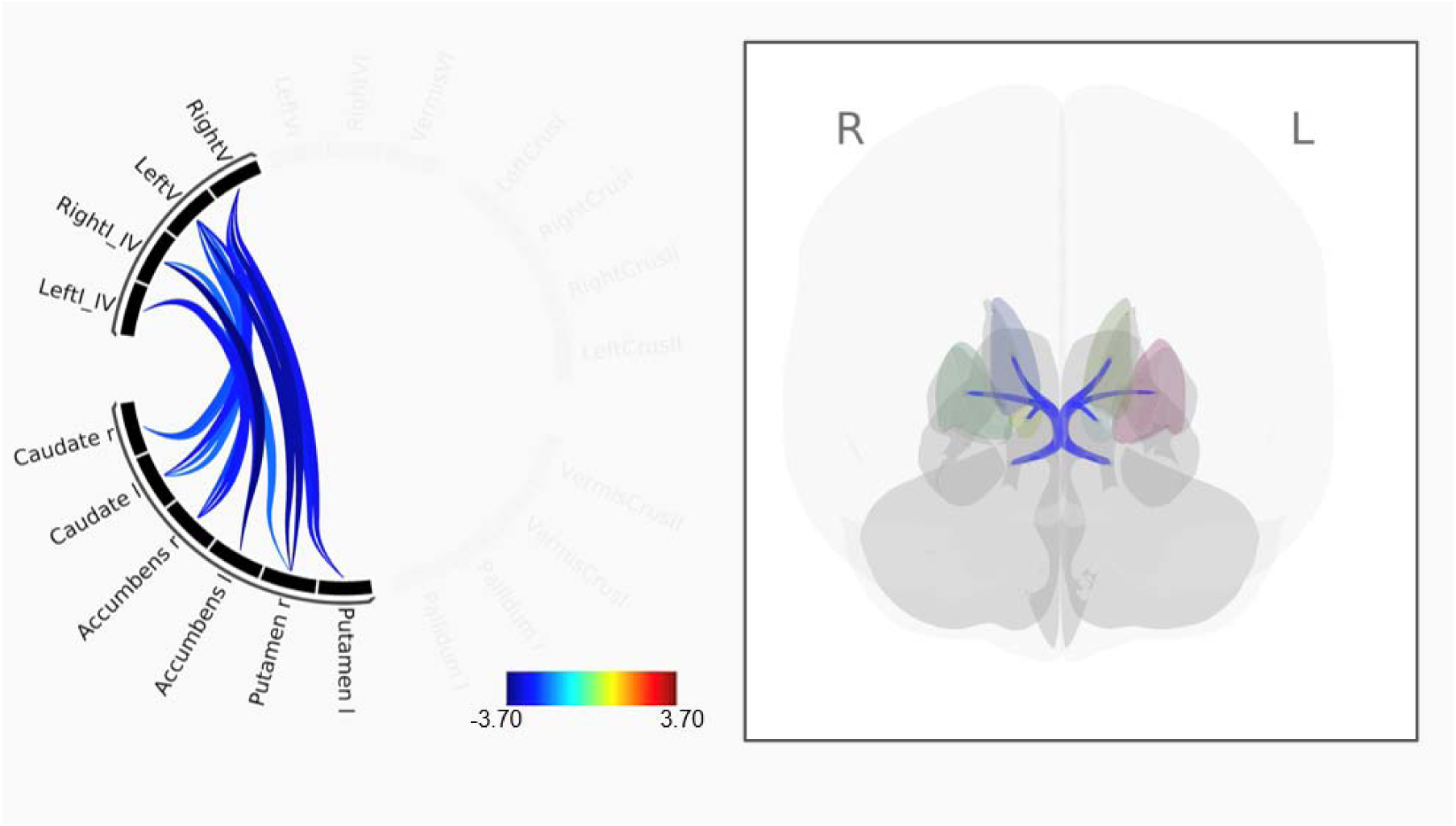
Age-related alterations in cerebellar–basal ganglia connectivity. Age is negatively associated with cerebello—basal ganglia connectivity across individuals with PD. Left: connectome ring showing lower connectivity (anticorrelations) with greater age between regions of the cerebellum and basal ganglia, consistent with past work. The inset (gray box) includes a 3D visualization of these associations between the cerebellum and basal ganglia. The color bar indicates the strength of the relationships between ROIs. Chord diagrams depict the magnitude and direction of age-related associations. All results are thresholded at *p < 0.05* and cluster-level *pFDR < 0.05* (MVPA omnibus test). PD: Parkinson’s disease.

**Table 3.** Region of Interest (ROI) to ROI association between age and connectivity adjusted for dopaminergic medication.

| Region 1 | Region 2 | Statistic | *p-unc* | *p-FDR* |
| --- | --- | --- | --- | --- |
| Left accumbens | Right I to IV | T(169) = -3.70 | 0.0001 | 0.0029 |
| Right putamen | Left V | T(169) = -3.42 | 0.0004 | 0.0045 |
| Right putamen | Right V | T(169) = -3.38 | 0.0005 | 0.0045 |
| Left accumbens | Left I to IV | T(169) = -2.89 | 0.0022 | 0.0203 |
| Left caudate | Right V | T(169) = -2.70 | 0.0038 | 0.0209 |
| Right accumbens | Right I to IV | T(169) = -2.85 | 0.0025 | 0.0249 |
| Left putamen | Right V | T(169) = -2.93 | 0.0019 | 0.0329 |
| Left putamen | Left V | T(169) = -2.64 | 0.0045 | 0.0329 |
| Right accumbens | Left I to IV | T(169) = -2.58 | 0.0054 | 0.0361 |
| Left caudate | Left V | T(169) = -2.14 | 0.0170 | 0.0363 |
| Right putamen | Left I to IV | T(169) = -1.98 | 0.0245 | 0.0377 |
| Left caudate | Right I to IV | T(169) = -1.98 | 0.0245 | 0.0409 |
| Right caudate | Right V | T(169) = -2.21 | 0.0142 | 0.0472 |
| Right caudate | Left V | T(169) = -2.11 | 0.0183 | 0.0506 |
| Right accumbens | Left V | T(169) = -2.26 | 0.0126 | 0.0570 |
| Right accumbens | Right V | T(169) = -1.94 | 0.0268 | 0.0767 |
| Left accumbens | Right V | T(169) = -1.84 | 0.0337 | 0.0857 |

Connectivity results were considered significant at a connection-level threshold of $p < 0.05$ and a cluster-level threshold of $pFDR < 0.05$ (MVPA omnibus test).

### Effect of disease duration

We next investigated whether disease duration can impact cerebellar functional connectivity in PD. When looking first at the ROI-ROI, longer disease duration was associated with robust positive connectivity between right and vermis lobule VI and the right pallidum, in addition to within cerebellar and basal ganglia connectivity (**Figure 3A**; **Table 4**). However, while there were no significant group differences when comparing cognitively normal PD participants relative to those with cognitive dysfunction, there are notable qualitative differences. Within the cognitively normal sample, there are several connections within both the basal ganglia and cerebellum, but these are far fewer in those with cognitive decline (**Figure 3C-D**, **Table 4**). When exploring these associations in whole-brain seed-to-voxel analyses, in cognitively normal PD, longer disease duration was robustly associated with stronger Lobule V – middle and inferior temporal gyrus connectivity, while there were negative associations with premotor and prefrontal regions (**Figure 3B**, **Table 5**). Furthermore, the group-level analyses showed that cognitively normal PD participants exhibited higher Crus I – left precentral gyrus connectivity compared to cognitive decline participants, even after controlling for disease duration (**Figure 4**) (coordinates: -28, -14, 52, cluster size=571 voxels, TFCE=966.85, peak p-FEW=.018). This pattern suggests that cerebello–cortical connectivity from Crus I to motor cortical regions, which is not typically a part of the Crus I network,^49^ may be a source of compensation in PD for those without cognitive decline. There were no associations with Crus I when looking at the full PD sample.

**Figure 3.**
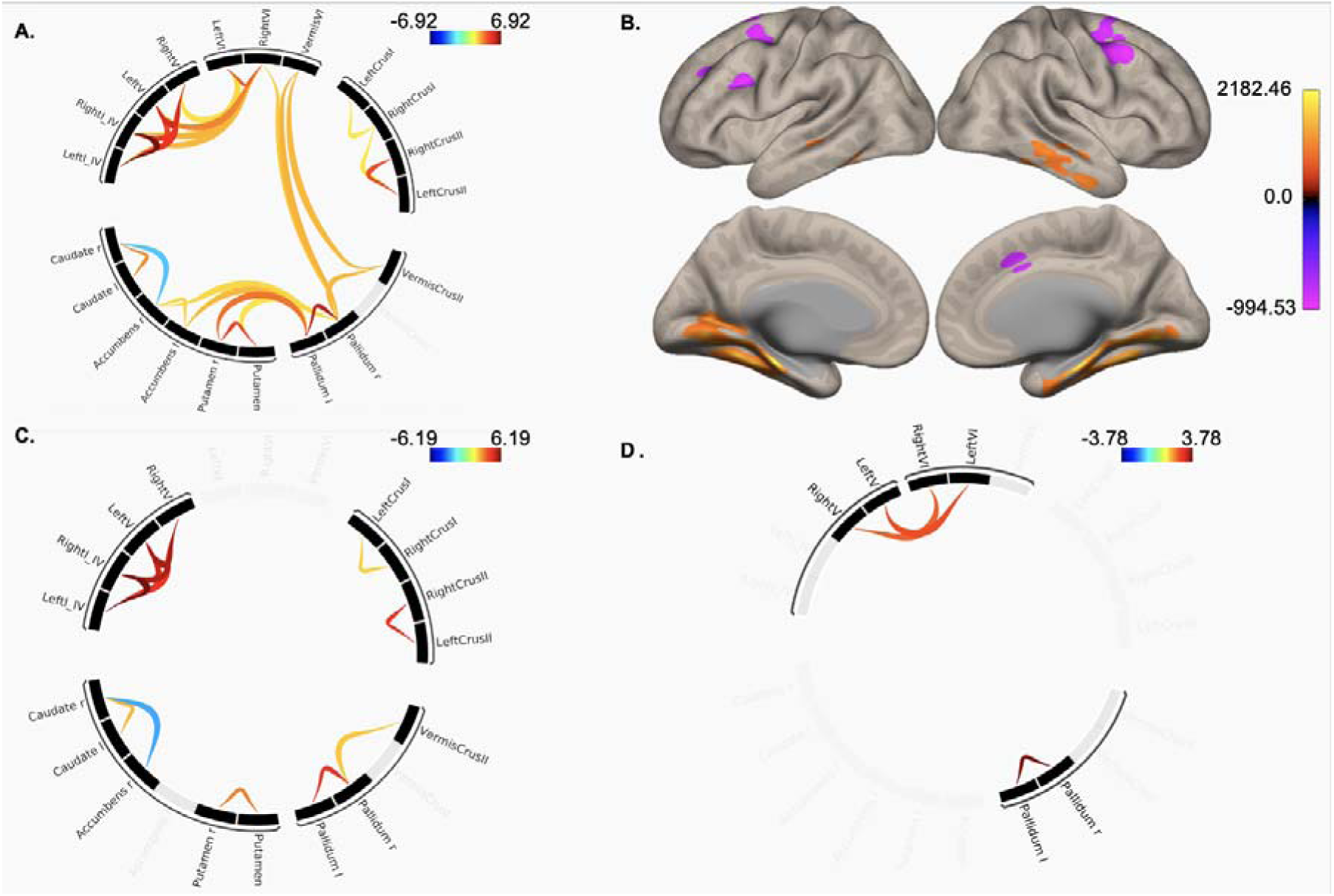
Disease duration effects on cerebellar–cortical connectivity. A. Associations between connectivity and disease duration in the entire PD sample largely demonstrating patterns of within structure (cerebellum and basal ganglia) connectivity as disease duration lengthens. B. Exploratory whole-brain analyses of Lobule V connectivity associations with disease duration. Connectivity with medial temporal lobe regions was increased with longer disease duration, while it was decreased with premotor and prefrontal areas. C. Connectome ring demonstrating associations between cerebello-basal ganglia connectivity and disease duration in cognitively normal PD only, and D. PD decline. There are no significant group differences between the cognitively normal and cognitive decline groups; however, qualitatively, the associations with disease course are fewer in the cognitive decline group. Color bars are indicative of connectivity strength. For panel B, numbers indicate TFCE values. Maps are thresholded at a connection-level *p < 0.05* and cluster-level *pFDR < 0.05* (MVPA omnibus test). PD: Parkinson’s disease.

**Figure 4.**
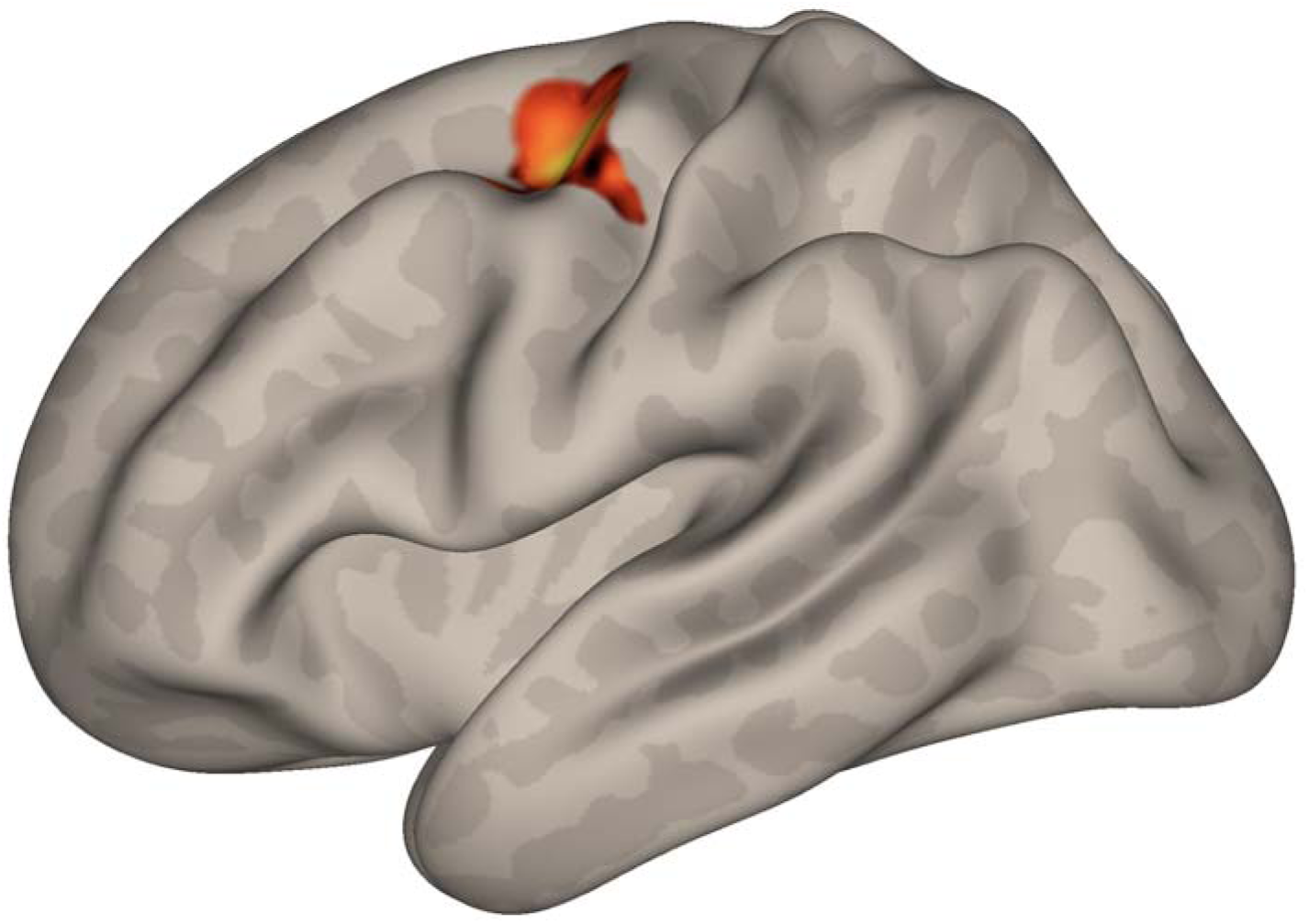
Cognitive-group differences in Crus I–precentral gyrus connectivity. Cognitively normal PD participants demonstrated higher Crus I–left precentral gyrus connectivity than PD-MCI and PDD participants, even after controlling for disease duration. This connectivity pattern aligns with preserved cerebellar–cortical support for cognition in early PD. Color bar indicates strength of connectivity as indexed by TFCE values. Results are thresholded at *p < 0.05* and cluster-level *pFDR < 0.05* (MVPA omnibus test). PD: Parkinson’s disease; PD-MCI: PD with mild cognitive impairment; PDD: Parkinson’s disease dementia.

**Table 4.** Region of Interest (ROI) to ROI effects of disease duration on connectivity adjusted for dopaminergic medication, with separate sections demonstrating for whole participants, cognitively normal PD, and PD with cognitive dysfunction.

| Group | Region 1 | Region 2 | Statistic | p-unc | p-FDR |
| --- | --- | --- | --- | --- | --- |
| **All PD** | Right I to IV | Left I to IV | T(169) = 6.92 | < 0.0001 | < 0.0001 |
|  | Right V | Left V | T(169) = 5.52 | < 0.0001 | < 0.0001 |
|  | Right V | Left I to IV | T(169) = 5.04 | < 0.0001 | < 0.0001 |
|  | Right V | Right I to IV | T(169) = 4.96 | < 0.0001 | < 0.0001 |
|  | Left V | Left I to IV | T(169) = 4.49 | < 0.0001 | 0.0001 |
|  | Left V | Right I to IV | T(169) = 4.44 | < 0.0001 | 0.0001 |
|  | Left pallidum | Right pallidum | T(169) = 5.94 | < 0.0001 | < 0.0001 |
|  | Right pallidum | Vermis Crus II | T(169) = 2.72 | 0.0073 | 0.0488 |
|  | Right putamen | Left putamen | T(169) = 4.50 | < 0.0001 | 0.0003 |
|  | Right caudate | Left caudate | T(169) = 3.28 | 0.0013 | 0.0250 |
|  | Right accumbens | Left accumbens | T(169) = 2.34 | 0.0202 | 0.1458 |
|  | Right caudate | Right accumbens | T(169) = -2.47 | 0.0146 | 0.1460 |
|  | Left Crus II | Right Crus II | T(169) = 4.38 | < 0.0001 | 0.0004 |
|  | Right Crus I | Left Crus I | T(169) = 2.04 | 0.0433 | 0.2167 |
|  | Left Crus II | Right Crus I | T(169) = 2.04 | 0.0426 | 0.2840 |
|  | Right VI | Left V | T(169) = 3.47 | 0.0006 | 0.0065 |
|  | Right VI | Right I to IV | T(169) = 2.78 | 0.0060 | 0.0401 |
|  | Right VI | Left I to IV | T(169) = 2.68 | 0.0081 | 0.0404 |
|  | Right VI | Right V | T(169) = 2.17 | 0.0312 | 0.1041 |
|  | Left VI | Left V | T(169) = 2.25 | 0.0260 | 0.1555 |
|  | Left VI | Right I to IV | T(169) = 2.17 | 0.0313 | 0.1555 |
|  | Vermis Crus II | Vermis VI | T(169) = 2.69 | 0.0079 | 0.0393 |
|  | Right pallidum | Vermis VI | T(169) = 2.76 | 0.0064 | 0.0488 |
|  | Right pallidum | Right VI | T(169) = 2.46 | 0.0151 | 0.0754 |
|  | Right VI | Left VI | T(169) = 3.93 | 0.0001 | 0.0025 |
|  | Right putamen | Left pallidum | T(169) = 3.75 | 0.0002 | 0.0025 |
|  | Right accumbens | Right pallidum | T(169) = 2.29 | 0.0231 | 0.1458 |
|  | Right accumbens | Left pallidum | T(169) = 2.20 | 0.0291 | 0.1458 |
|  | Left accumbens | Left pallidum | T(169) = 2.68 | 0.0081 | 0.1615 |
|  | Left accumbens | Right pallidum | T(169) = 2.21 | 0.0283 | 0.1886 |
|  | Left putamen | Right pallidum | T(169) = 2.12 | 0.0357 | 0.2377 |
| **PD: cognitively normal** | Right I to IV | Left I to IV | T(102) = 6.19 | < 0.0001 | < 0.0001 |
|  | Right V | Left V | T(102) = 5.55 | < 0.0001 | < 0.0001 |
|  | Right V | Right I to IV | T(102) = 5.46 | < 0.0001 | < 0.0001 |
|  | Left V | Right I to IV | T(102) = 5.14 | < 0.0001 | < 0.0001 |
|  | Right V | Left I to IV | T(102) = 4.49 | < 0.0001 | 0.0001 |
|  | Left V | Left I to IV | T(102) = 4.45 | < 0.0001 | 0.0001 |
|  | Left pallidum | Right pallidum | T(102) = 4.64 | < 0.0001 | 0.0002 |
|  | Right pallidum | Vermis Crus II | T(102) = 2.22 | 0.0290 | 0.2897 |
|  | Right putamen | Left putamen | T(102) = 3.15 | 0.0022 | 0.0296 |
|  | Right caudate | Right accumbens | T(102) = -2.61 | 0.0104 | 0.1740 |
|  | Right caudate | Left caudate | T(102) = 2.42 | 0.0174 | 0.1740 |
|  | Left Crus II | Right Crus II | T(102) = 4.34 | < 0.0001 | 0.0007 |
|  | Right Crus I | Left Crus I | T(102) = 2.14 | 0.0350 | 0.2334 |
| **PD: cognitive decline** | Left VI | Left V | T(64) = 2.39 | 0.0200 | 0.1331 |
|  | Right VI | Right V | T(64) = 2.22 | 0.0299 | 0.1990 |
|  | Left Pallidum | Right pallidum | T(64) = 3.78 | 0.0004 | 0.0070 |

Connectivity results were considered significant at a connection-level threshold of $p < 0.05$ and a cluster-level threshold of $pFDR < 0.05$ (MVPA omnibus test).

**Table 5.** Lobule V whole brain connectivity associated with disease duration in Parkinson’s disease adjusted for dopaminergic medication.

| Region | Cluster (x,y,z) | Size | Peaks | TFCE | peak p-FDR |
| --- | --- | --- | --- | --- | --- |
| Vermis IV to VI | -06 -36 -24 | 7880 | 60 | 3120.01 | < 0.0001 |
| Right precentral gyrus | +44 +04 +42 | 702 | 10 | 1021.96 | 0.0112 |
| Left middle frontal gyrus | -28 +08 +34 | 437 | 9 | 995.66 | 0.0121 |
| Left superior frontal gyrus | -16 +04 +58 | 132 | 3 | 922.70 | 0.0162 |
| Medial anterior cingulate gyrus | +06 +14 +36 | 100 | 1 | 866.40 | 0.0179 |
| Left frontal pole | -28 +40 +28 | 45 | 1 | 864.89 | 0.0179 |
| Right middle frontal gyrus | +30 +16 +24 | 1 | 1 | 844.00 | 0.0189 |
| Right inferior frontal gyrus/frontal operculum | +32 +16 +20 | 2 | 1 | 841.79 | 0.0189 |

Connectivity results were considered significant at a connection-level threshold of $p < 0.05$ and a cluster-level threshold of $pFDR < 0.05$ (MVPA omnibus test).

### Association with motor symptom severity

We additionally examined whether motor symptom severity, measured by the MDS-UPDRS Part III, was associated with cerebellar functional connectivity. Across all analyses, no significant associations emerged between motor severity and any connectivity measures, regardless of whether dopaminergic medication status was controlled. These results indicate that clinical motor symptom burden did not systematically relate to cerebellar network alterations within this cohort.

## Discussion

Our findings provide strong support for the compensatory role of the cerebellum in PD even in the presence of cognitive decline. Recent conceptualizations of the cerebellum in aging have suggested that it provides critical scaffolding for cortical function in aging, in the face of age-related structural and functional declines.^20,50,51^ This parallels the concept of cerebellar reserve wherein relatively preserved cerebellar resources may compensate in the face of neurodegenerative disease.^24,52^ Our present study has provided further support for this framework in PD. Enhanced cerebello-precentral gyrus and cerebello-pallidal connectivity in cognitively normal PD suggests that the cerebellum is recruited to scaffold cognition during early disease stages.^19^ The concurrent involvement of motor (precentral gyrus, pallidum) and association cortices underscores that cerebellar compensation in PD is not confined to “cognitive” circuits but instead spans integrated motor–cognitive networks that support goal-directed behavior. These results are consistent with theories of cognitive reserve and neural scaffolding,^24,52–54^ which posit that the brain adapts to pathology by engaging alternative networks. Importantly, we showed that purportedly compensatory cerebellar connectivity relates to disease duration and cognitive status, even in the context of widespread age-related declines in cerebello–basal ganglia coupling. In addition, the absence of significant associations between MDS-UPDRS motor severity and cerebellar connectivity suggests that these compensatory processes are not directly linked to clinical motor burden, but rather reflect network-level adaptations that may operate independently of motor symptom severity.

With longer disease duration, however, compensatory mechanisms decline, as evidenced by reduced cerebello-cortical connectivity in association networks. This may suggest that cerebellar reserve has limits, and once exceeded, patients progress more rapidly toward dementia; however, longitudinal investigations are critical for investigating this more directly. These findings underscore the dynamic nature of cerebellar contributions, with compensatory network engagement evident in cognitively normal PD but diminished as cognitive dysfunction emerges. Importantly, our disease-duration analyses also revealed that in cognitively normal PD, Lobule V connectivity with middle and inferior temporal cortex was higher with longer disease duration, consistent with targeted, region-specific recruitment that may help sustain cognitive function despite advancing pathology. These regions are outside the canonical Lobule V motor network.^47–49^ In contrast, only a small cerebellar cluster showed disease-duration effects in the cognitive-decline group, further supporting the notion that compensatory capacity diminishes as cognitive impairment emerges.

Our findings also resonate with broader work on aging-related cerebellar and dopaminergic decline. The spatial distribution and magnitude of the anti-correlations in our study findings closely mirrored previously reported cerebello–basal ganglia patterns observed in older adults, with aging in PD characterized by reduced synchrony between cerebellar and striatal nodes.^25^ Seidler and colleagues reviewed evidence showing that older adults experience structural atrophy of the cerebellum alongside degeneration of dopaminergic systems, especially within the nigrostriatal pathway.^20^ These changes contribute to motor slowing, postural instability, and reduced coordination, even in otherwise healthy aging. Importantly, such dopaminergic denervation places the aging brain on a continuum with preclinical Parkinson’s disease.^20^ The parallel vulnerability of both cerebellar and basal ganglia circuits suggests that the altered cerebellar connectivity we observed in Parkinson’s disease may, in part, reflect an amplification of mechanisms already present in aging. This convergence highlights the cerebellum’s integral role in compensatory processes and circuits but also underscores its susceptibility when dopaminergic inputs are diminished. Thus, our study extends prior work by demonstrating how basal ganglia–cerebellar interactions may shape the trajectory from normative aging to Parkinsonian pathology, with and without cognitive decline. Of note, our medication-controlled analyses revealed reduced Lobule V – M1 connectivity across PD subgroups, suggesting that dopaminergic medication is associated with cerebello–motor coupling; however, there were no group differences, indicating broadly similar dopaminergic medication sensitivity across cognitive subtypes.

Clinically, the observation that cerebellar connectivity shows disease-duration–dependent and Crus I–motor cortical coupling in cognitively normal Parkinson’s disease, but not in those with cognitive decline, suggests that cerebellar networks may reflect compensatory capacity that is not captured by standard clinical scales, supporting their potential relevance as imaging biomarkers and neuromodulatory targets (e.g., transcranial magnetic stimulation) to serve as a promising avenue to sustain compensatory capacity. However, several limitations temper these implications. The cross-sectional, resting-state fMRI design yields correlational measures and precludes inference about longitudinal trajectories of cerebellar compensation or decline, underscoring the need for replication in prospective cohorts integrating task-based and structural imaging. Clinical classification schemes, including MoCA cutoffs and MDS-UPDRS severity categories, may not fully capture the complexity of cognitive and motor progression in PD, potentially obscuring more subtle brain–behavior relationships and contributing to the limited connectivity differences observed across UPDRS-based severity. Finally, because participants were grouped primarily by cognitive status rather than detailed motor phenotypes, future work should incorporate more granular composite clinical indices that integrate both motor and non-motor progression to better disentangle domain-specific versus shared cerebellar compensatory mechanisms over time.

## Conclusion

In summary, cerebellar functional connectivity shows dynamic, region-specific alterations across the cognitive stages of Parkinson’s disease. Several enhanced connectivity patterns in those that are cognitively normal, and in an earlier more intact cognitive stage are suggestive of compensatory support. These results highlight cerebello-basal ganglia and –cortical connectivity as a promising neuroimaging biomarker of cognitive disease progression and treatment response, and underscore the need to determine how preserved or strengthened cerebellar pathways can be harnessed for therapeutic development, including targeted neuromodulation.

## Data Availability

All data produced in the present study are available upon reasonable request to the authors

https://www.ppmi-info.org/

## Author’s Role

Lin CR: study design and execution, data analysis, initial manuscript writing and critical revision of the manuscript for intellectual content;

Magalhães TNC: data retrieval and organization, data analysis, data interpretation;

Yonce SS: data retrieval and organization;

Rampalli I: data retrieval and organization;

Mahabir R: study execution, IRB writing and revision;

Bernard JA: study design and execution, data analysis and interpretation, and critical revision of the manuscript for intellectual content

## Acknowledgement

Statistical analysis codes used to perform the analyses in this article are shared on Zenodo [10.5281/zenodo.19981833]. PPMI – a public-private partnership – is funded by the Michael J. Fox Foundation for Parkinson’s Research and funding partners, including AbbVie, Alamar Biosciences, Aligning Science Across Parkinson’s (ASAP), Arrowhead Pharma, Arvinas, AskBio, BIAL, BioArctic, Biohaven, BlueRock Therapeutics, Bristol Myers Squibb, Calico Labs, Capsida Biotherapeutics, Critical Path Institute, DaCapo Brainscience, Denali, Edmond J. Safra Foundation, Eli Lilly, Gain Therapeutics, GE Healthcare, Genentech, GSK, Insitro, Johnson & Johnson Innovative Medicine, Lundbeck, Merck, Neumora, Neuron23, Novartis, Olink, Regeneron, Roche, Sanofi, Tenvie, UCB, Vanqua Bio, Voyager Therapeutics, The Weston Family Foundation. The PPMI author list can be found in the supplemental file.

